# Timing of decompressive craniectomy and in-hospital mortality in children following severe traumatic brain injury using the German Hospital Database

**DOI:** 10.1101/2024.11.13.24317240

**Authors:** Rayan Hojeij, Pia Brensing, Bernd Kowall, Andreas Stang, Michael Nonnemacher, Ursula Felderhoff-Müser, Philipp Dammann, Marcel Dudda, Christian Dohna-Schwake, Nora Bruns

**Author notes:** **Corresponding Author:** Rayan Hojeij, Address: Universitätsklinikum Essen, Hufelandstraße 55, 45147, Essenm, Telephone Number: +49/201-2451 or -7192.

## Abstract

**Introduction:** Decompressive craniectomy (DC) is a last tier to control refractory intracranial pressure elevation in children with severe traumatic brain injury (sTBI), but the optimal timing is unknown. This study aimed to describe the association between timing of DC and in-hospital mortality in children with sTBI in Germany.

**Methods:** A retrospective cohort study of the German national hospital discharge database (2016-2022) was conducted for cases < 18 years. Children undergoing DC following sTBI were extracted. Time from admission to DC were calculated as complete hours and data were compared between early (time to DC ≤ 2 hours) and late DC (> 2 hours). Hierarchical logistic regression models evaluated the association of DC timing with in-hospital mortality, functional outcomes (Pediatric Complex Chronic Conditions (PCCC) ≥2)), poor outcome (composite outcome of death or PCCC≥2), length of hospital stay, days on mechanical ventilation (MV) and coding of seizures.

**Results:** Among 13,492,821 children hospitalized, 9,495 had sTBI. DC was performed in 598 cases with time to decompression ranging from 0 to 27 days. More than half of DCs (54.8%) were performed within the first two hours after admission. 164 (27.6%) deaths occurred, with a median time from admission to death of 2 days. Early DC had a higher case-fatality (37.4%) compared to late DC (15.8%), with higher odds of death (adjusted odds ratio [OR] 2.89; 95% confidence interval [95%CI] 1.43-5.85) and poor outcome (OR 1.22; 95% CI 0.71-2.21). However, in survivors, early DC was associated with a shorter duration of MV. No differences in functional outcomes were associated with the timing of DC.

**Conclusion:** Children undergoing early DC exhibited a higher risk of case fatality and poor outcome, alongside with less days on MV in survivors.

**Key points:** *Question:* Is there an association between the timing of decompressive craniectomy (DC) and in-hospital mortality in children with severe traumatic brain injury (sTBI)?

*Finding:* Early DC (≤ 2 hours after admission) was associated with higher case fatality and poor outcomes but fewer days on mechanical ventilation in survivors compared to late DC (> 2 hours).

*Meaning:* The timing of DC may impact survival and recovery outcomes in pediatric sTBI, suggesting the need to balance early intervention with potential risks.

## Introduction

Decompressive craniectomy (DC) has emerged as a rescue measure to relieve refractory intracranial hypertension and prevent secondary cerebral ischemia or herniation in children with severe traumatic brain injury (sTBI). Primary DC, performed in the initial phase when swelling occurs or is anticipated, is often carried out concurrently with hematoma evacuation following severe head trauma [1]. On the other hand, secondary DC serves as a last-tier option to treat refractory elevation of intracranial pressure (ICP) despite medical management (MM) during the clinical course [2].

Very few randomized controlled trials (RCT) have evaluated the effect of DC on outcomes in adults and children with refractory or persistent ICP elevation. The only pediatric RCT that included 27 children found that DC performed at a median time of 17 hours after admission was associated with good outcomes and significant ICP reduction compared to the MM group [3]. The DECRA trial was conducted among adults with severe diffuse traumatic brain injury and ICP elevation above 20 mmHg for more than 15 min for a one-hour period within the first 72 hours after injury. Patients were randomized to either bifronto-temporo-parietal DC or standard care [4, 5]. Early DC was associated with poorer functional outcomes at 6 months of follow-up[5], and higher vegetative state in survivors at 12 months of follow-up[4]. In the RESCUEicp trial, patients ≥ 10 years were randomized to undergo DC (as a last tier) after optimized MM, or to receive ongoing medical care if the ICP levels remained above 25 mmHg for 1 to 12 hours [6]. Six-month outcomes yielded lower lethality but higher frequencies of vegetative state than the medical care group. Due to these inconsistent trials regarding the timing of decompression and their contradicting results, recommendations regarding DC remain rather imprecise, ultimately leaving the decision to decompress or not at the discretion of the treating medical team.

With respect to the optimum timing of DC in pediatric TBI, observational studies comparing the outcomes of “early” versus “late” DC have produced varying results. One systematic review (including case series studies)[7], and two retrospective studies reported that early DC, defined as time to DC of less than 12 and 24 hours respectively, is beneficial in cases of refractory ICP, reducing morbidity and mortality in children[8, 9], whereas Nagy et al. found no association between timing of DC and outcomes in children [10]. Despite these contradictory findings on the general relationship between timing and outcomes, the literature lacks a universal definition for “early” or “late” DC, as studies used varying cut-off times to define groups, ranging from 2 to 24 hours. Further, the timing of DC is influenced by factors such as injury severity, neurological status, individual patient characteristics, health care system infrastructure, and the neurosurgeon’s decision.

The aim of this study was to assess the timing of DC and associated outcomes in the German hospital dataset (GHD) in pediatric cases with sTBI. Using routine health care data on hospitalizations of the entire country, this study provides information on timing aspects of DC in pediatric TBI care.

## Methodology

### 1. Materials and methods

#### 2.1. Study Design

We conducted a retrospective cohort study using patient data from the German Hospital Dataset (GHD) to investigate the association between timing of DC and in-hospital lethality in pediatric cases with severe traumatic brain injury (sTBI). The comprehensive nature of the study and dataset allows an exhaustive analysis of a large dataset, encompassing the entire public hospitals across the country.

German hospitals have been reimbursed based on diagnosis related groups (DRGs) since 2004. According to §21 KHEntgG, German hospitals are required by law to share data on all hospital admissions with the hospital payment system (InEK). After passing plausibility checks, these data are anonymized and sent to the Federal Statistical Office (FSO). As these hospitalization data are mandatory for reimbursement, hospitals are strongly incentivized to provide complete data sets. Detailed information on the structure of the DRG dataset is available at the FSO and further details on the process of data access at https://www.forschungsdatenzentrum.de/en/health/drg.

#### 2.2. Case selection

Inclusion criteria were cases < 18 years of age discharged from public hospitals in Germany between 2016 and 2022 with sTBI who underwent DC. Selected cases had TBI as primary discharge diagnosis (ICD-10 code: S06) identified via codes of the International-Classification of Disease, 10^th^ edition, German modification (ICD-10-GM). We defined sTBI as an Abbreviated Injury Scale (AIS) of the head ≥ 3. Patients who underwent decompressive craniectomy were identified and selected via operation and procedure (OPS) codes (5-0120, 5-0101, 5-0104) (Table 1).

**Table 1:** Children’s characteristics stratified by the timing of surgery after hospital admission for severe traumatic brain injury in Germany, 2016-2022.

|  | Total (N=589),<br>No.(%) | Early DC (n=323),<br>No.(%) | Late DC (n=266),<br>No. (%) |
| --- | --- | --- | --- |
| Age, median (IQR) | 12 (4-16) | 12 (3-16) | 11 (5-16) |
| Age categories |  |  |  |
| 0-5 | 171 (29.0) | 96 (29.7) | 75 (28.2) |
| 6-12 | 121 (20.5) | 60 (18.6) | 61 (22.9) |
| 13-17 | 297 (50.4) | 167 (51.7) | 130 (48.9) |
| Sex, male | 395 (67.1) | 220 (68.1) | 175 (65.8) |
| LOS in days, median (IQR) | 17 (5-29) | 11 (2-23) | 21.5 (9-32) |
| LOS in survivors, median (IQR) | 22 (13-35) | 19 (11-33) | 24 (16-35) |
| Transferred cases | 50 (8.4) | 27 (8.3) | 23 (8.6) |
| In-hospital mortality | 163 (27.7) | 121 (37.5) | 42 (15.8) |
| 14-days In-hospital mortality, | 155 (26.3) | 116 (35.9) | 39 (14.7) |
| Time to death after DC,<br>median (IQR) | 2 (1-6) | 2 (1-4) | 4 (1-5) |
| Mechanical ventilation (days),<br>median (IQR) | 7.5 (2.5-15.8) | 5 (1.7-12) | 10.8 (5.9-19) |
| Serious extracranial injuries (AIS≥ 3) | | | |
| Thorax | 195 (33.1) | 118 (36.5) | 77 (28.9) |
| Abdomen | 28 (4.7) | 21 (6.5) | 7(2.6) |
| Spine | 11 (1.9) | 6 (1.9) | 5 (1.8) |
| Type of head injury |  |  |  |
| EH (Epidural) | 166 (28.2) | 92 (28.5) | 74 (27.8) |
| SH (Subdural) | 411 (69.8) | 231 (71.5) | 180 (67.7) |
| SAH (Subarachnoid) | 223 (37.9) | 116 (35.9) | 107 (40.2) |
| Brain oedema | 382 (64.8) | 210 (65.0) | 172 (64.7) |
| Complications |  |  |  |
| IHCA | 67 (11.4) | 35 (10.8) | 32 (12.0) |
| OHCA | 20 (3.4) | 11 (3.4) | 9 (3.4) |
| Coma | 184 (31.2) | 112 (34.7) | 72 (27.1) |
| Seizure | 73 (12.4) | 34 (10.5) | 39 (14.7) |
| Epilepsy | 18 (3.0) | 5 (1.5) | 13 (4.9) |
| DIC | 30 (5.0) | 24 (7.4) | 6 (2.3) |
| PCCC in survivors, median (IQR) | 2 (1-3) | 2 (1-2) | 2 (0-3) |
| Evacuation of hematoma | 373 (63.3) | 233 (72.1) | 140 (52.6) |
| EVD | 126 (21.4) | 56 (17.3) | 70 (26.3) |
| Preoperative | xx | xx | 41/70 (58.6) |
| Intraoperative | 48 (38.1) | 32/56 (57.1) | 16/70 (22.8) |
| Postoperative | xx | xx | 13/70 (18.6) |
| Time to EVD (days), median (IQR) | NA | 2 (1-29.5) | 13 (1-73) |
| ICP monitoring | 307 (52.1) | 153 (47.4) | 154 (57.9) |
| Preoperative | 127 (41.3) | 7/153 (4.5) | 120/154 (77.9) |
| Intraoperative | 159 (51.8) | 132/153 (86.3) | 27/154 (17.5) |
| Postoperative | 21 (6.8) | 14/153 (9.2) | 7/154 (4.6) |
| Time to ICP (minutes), median (IQR) | NA | 67 (38-105) | 120 (65-428) |
| POFI, median (IQR) | 0 (0-0) | 0 (0-0) | 0 (0-0) |
| ISS, median (IQR) | 13 (9-22) | 13 (9-22) | 13 (9-19) |
| Single ICISS, median (IQR) | 0.67(0.56-0.77) | 0.62 (0.50-0.77) | 0.68 (0.56-0.77) |
| Multiple ICISS, median (IQR) | 0.11(0.02-0.32) | 0.11 (0.02-0.31) | 0.13 (0.03-0.34) |

Abbreviations: DC= Decompressive craniectomy LOS= length of stay, IHCA= Intrahospital cardiac arrest, OHCA= Out of hospital cardiac arrest, DIC= Disseminated intravascular coagulation, PCCC= Pediatric Complex Chronic Conditions, EVD=Extra ventricular drain, ICP= Intracranial pressure, POFI= Postoperative Organ Failure Index, ISS= Injury Severity Score , ICISS= International Classification Injury Severity Score, IQR= Interquartile range, xx= Censored data, NA=not available.

#### 2.3. Data Extraction

We extracted data from the GHD, including patient demographics, type of injury, complications, survival status at discharge and length of stay (LOS). Time and date of admission, discharge, surgery for DC and ICP monitoring were extracted from the database and time from admission until surgery/placement of ICP monitor or EVD were computed.

The following type of head injury codes were extracted: traumatic cerebral oedema (ICD-10-GM: S06.1), traumatic subdural hemorrhage SH (ICD-10-GM: S06.5), traumatic epidural hemorrhage EH (ICD-10-GM: S06.4) and traumatic subarachnoid hemorrhage SAH (ICD-10-GM: S06.6).

Injury severity was quantified as previously described based on the AIS, injury severity score (ISS) and using a validated ICD based injury severity score (ICISS) [11]. These scores were used to categorize injuries and to adjust for injury severity. The ICISS is empirically derived by calculating survival proportions (or survival risk ratios SRR) for each injury diagnosis code. The ICISS was used to estimate injury severity using SRR. A lower SRR value indicates higher risk of death Single-ICISS was extracted as the single worst injury (= lowest value) from all trauma-related ICD codes for each case, while the multiplicative ICISS accounted for the multiple injuries by multiplying the assigned values of all trauma-related ICD codes of each case.

The Pediatric Complex Chronic Conditions (PCCC) Classification was used to assess the functional outcome at discharge with minor modifications that were necessary due to the inherent structure of the GHD [12] (eTable 1). The PCCC was designed to identify and quantify conditions in children that are likely to persist for at least one year. Organ dysfunction was assessed and quantified by summing up extracted binary scores that indicate organ dysfunction (eTable 2).

#### 2.4. Timing of surgery

Time from admission to DC was rounded to completed hours and categorized cases into two groups: early (within the first 2 completed hours after admission) and late (performed more than 2 hours post admission). This categorization was primarily data-driven, as there is no commonly accepted definition of early or late DC. The median number of hours to DC after admission was used as an indicator for the categorization.

#### 2.5. Outcomes measures

The primary outcome was in-hospital death following DC. Secondary outcomes included a poor functional outcome, defined as PCCC ≥ 2, proposed by Simon et al. as significant chronic conditions affecting body systems that are expected to last at least a year[13].This cut-off has also been used in other pediatric studies. Other secondary outcomes were LOS, duration of mechanical ventilation in days, coding of seizures, and poor outcome, defined as a composite outcome of death or PCCC ≥ 2.

#### 2.6. Missing data

There were no missing data on age, main diagnosis, LOS, or survival at discharge. In our analyses, we had to assume that an ICD-code or OPS code that was not documented meant that the diagnosis was not present, or the procedures was not done, respectively.

We nevertheless assume that extensively reimbursed procedures such as DC are carefully coded, which justifies our selection of these codes. In addition, 9 patients with missing information on time or date of admission, discharge or DC were removed from the final analysis.

#### 2.7. Statistical Analysis

Descriptive statistics were used to summarize demographic and clinical characteristics within each group. Continuous variables were described using median and interquartiles (Q1-Q3), while count data were presented as frequencies and percentages.

Characteristics associated with time to DC were described. Age, sex, type of head injury, coma, severity of the injury and organ dysfunction were considered as potential confounders based on the theory of directed acyclic graphs [14, 15], that are recommended for empirical pediatric and critical care research [16, 17] (eFigure 1).

Hierarchical logistic regression models were used to estimate the odds of outcomes associated with the timing of the intervention (early versus late DC). This method controls for the clustering of cases within centres (identified via the institutional identifier) [18, 19]. To account for clustering, institutional identifiers were included as a random effect in the model. Kaplan-Meier plots were generated to illustrate 14-day all-cause case fatality based on different time intervals to DC, with time starting from the date of DC. This ensures that the time-to-event analysis accurately reflects the post-DC period. We performed a sensitivity analysis of our primary outcome using the time ≤1 hour to surgery after admission to categorize early and late DC. All calculations and analyses were performed using SAS release 9.4 and SAS Enterprise Guide 7.3 (SAS Institute, Cary, North Carolina, USA).

## Results

Between 2016 and 2022, a total of 13,469,821 pediatric cases were discharged from public hospitals across Germany. Among these cases, 525,360 received a primary discharge diagnosis of TBI, 9,495 of which were severe TBI cases with AIS head ≥3. 598 (6.3%) cases underwent DC during their hospital stay, 323 (54.8%) within the first two hours and 443 (75.2%) within the first 24 hours of hospitalization (Fig. 1).

**Fig. 1:**
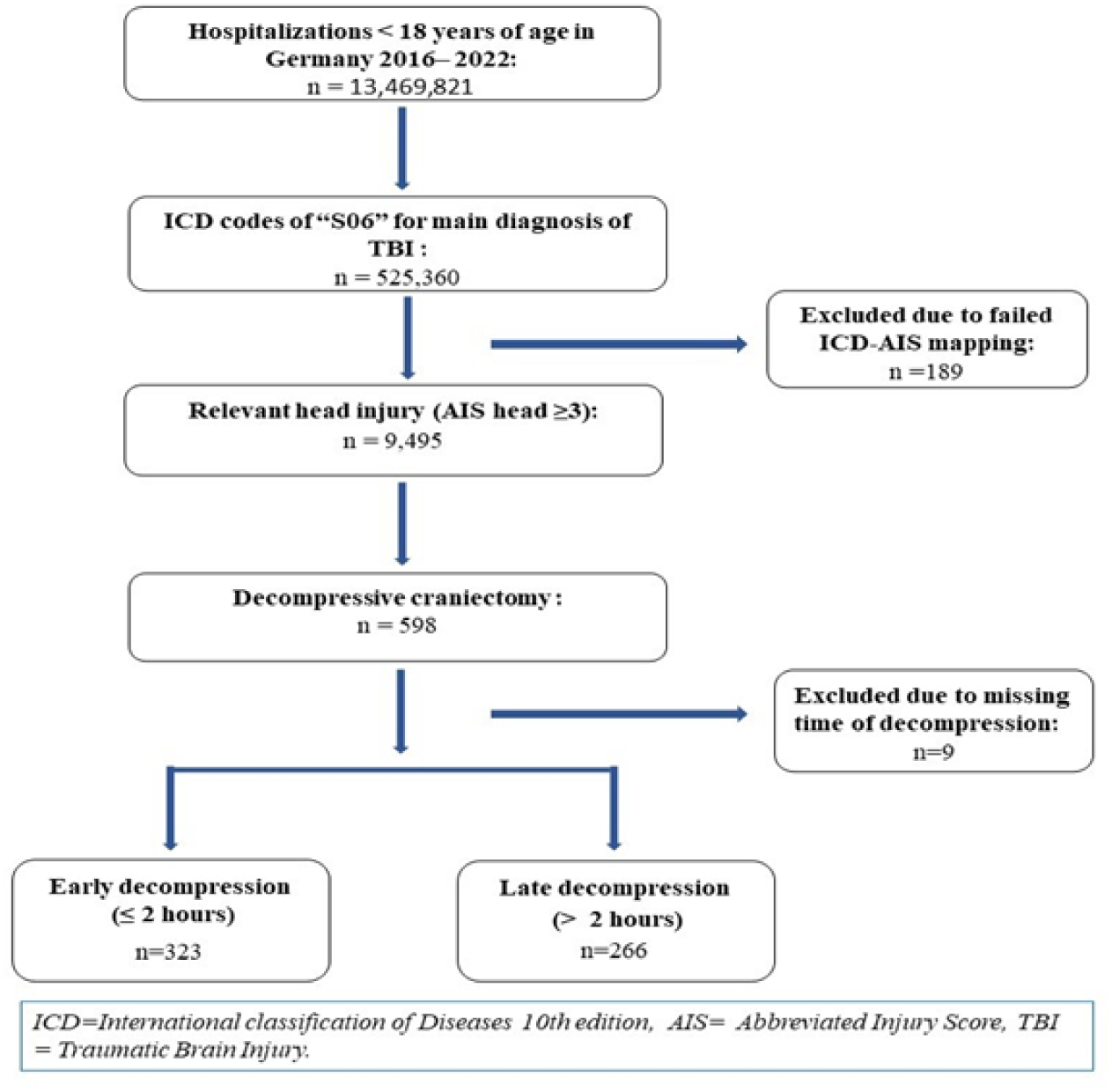
Flowchart of included traumatic brain injury cases aged 0-17 years in Germany, 2016-2022

The cases that received DC were predominantly male (67.1%) with a median age of 12 years (IQR 4 – 16) and a median hospital stay of 17 days. 27 % of cases died in hospital with a median time from admission to death of 2 days. The majority of DC cases involved brain edema and subdural hemorrhage, affecting 64.9% and 69.9 %, of patients, reported as separate findings respectively. One third of the cases were in coma (31.3%), and more than half received invasive ICP monitoring (51.8%).

Most DCs (54.8%) were performed within two hours of hospital admission, with numbers declining over the first 24 hours of admission (Fig. 2).

**Fig. 2:**
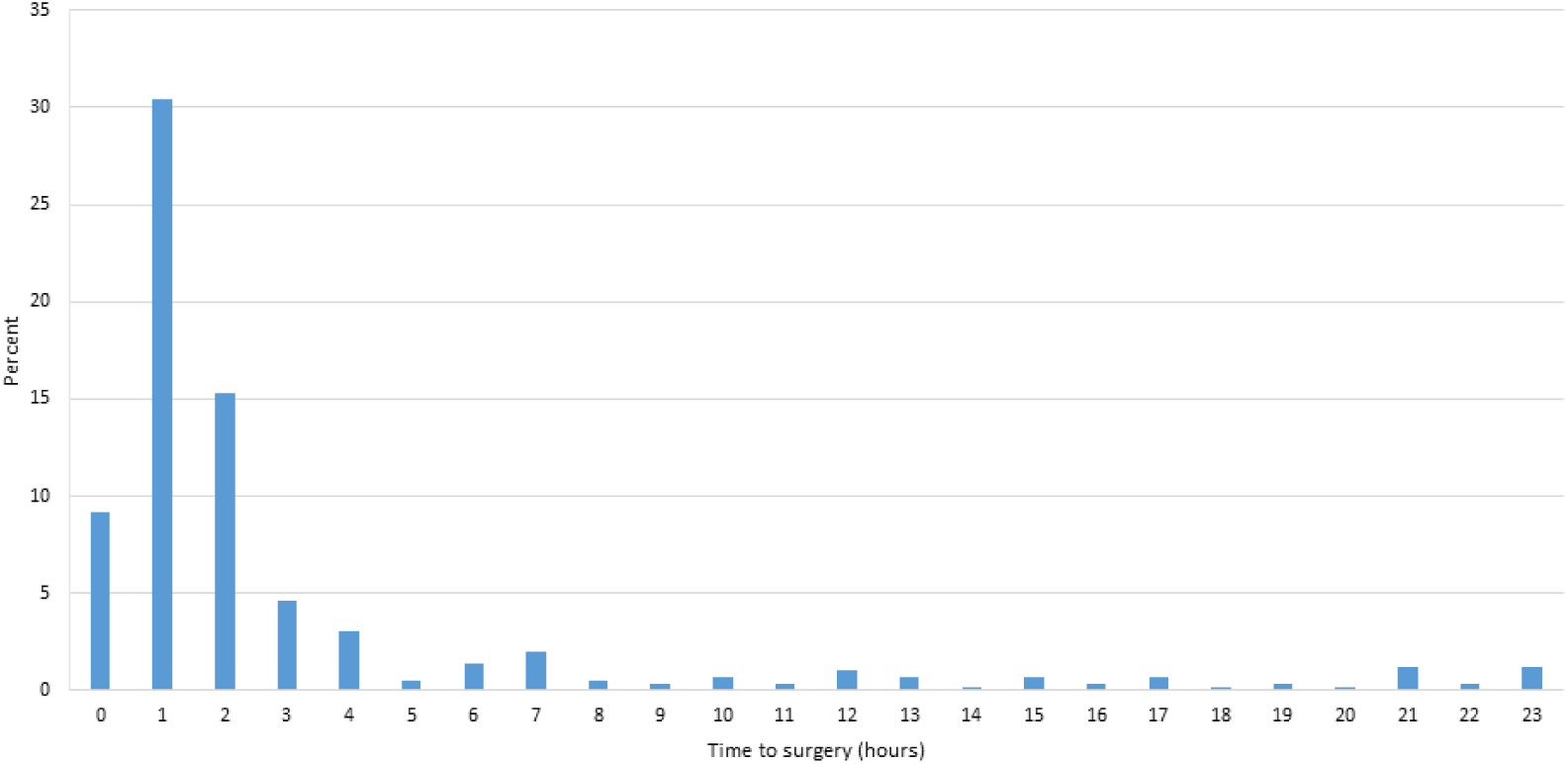
Distribution of time within the initial 24 Hours from admission to decompressive craniectomy surgery. *Hours are shown as complete hours. This graph represents 75% of the patients who underwent DC surgery in the first 24 h of admission.

Characteristics of patients receiving early, and late DC were largely similar (Table 1). Differences were observed for lethality (37.5% in early DC vs. 15.8% in late DC) and hematoma evacuation (72.0 % vs. 52.6%). A shorter median hospital stay of 11 days was observed in the early DC group compared to 22 days after late DC. While half of the cases received ICP monitoring in both the early and late DC group, 77.9% of the ICP monitoring in the late DC group was performed before DC. The median time from admission to ICP monitoring in the early DC group was 67 minutes, with 86% of ICP monitors being inserted during the same surgery in which the DC was performed. Similarly, we observed that 58.6% of those who received EVD were performed before the DC surgery in the late DC group, where 57.1% in the early DC group had EVD intraoperatively. No other clinically relevant differences between the groups were observed (Table 1).

The cumulative case fatality after DC based on time to DC after admission in hours is shown in Fig. 3 Postoperative cumulative mortality was higher when the time to craniectomy was within the first hour of admission after injury relative to longer hours (2 or 3 hours and more).

**Fig. 3.**
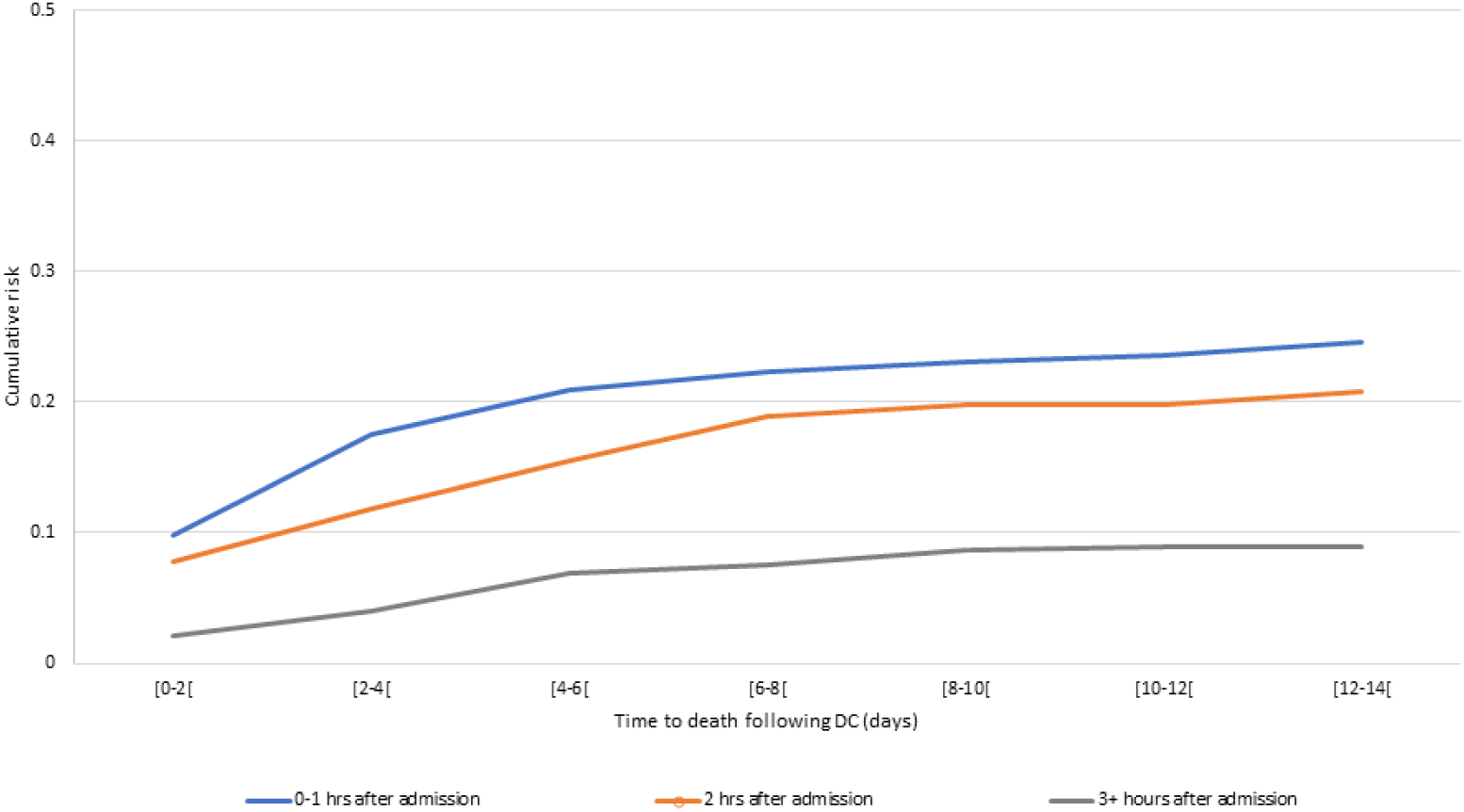
Cumulative case fatality following time to DC in hours by time to death within 14 days of hospitalization after the DC surgery. *Days and hours are shown as complete days/hours

Logistic regression showed higher adjusted odds for death (odds ratio (OR) 2.89 (95% CI: 1.43-5.85) and composite outcome (1.22 (0.71-2.21)) in early versus late DC groups (Figure 3). No differences were observed between early and late DC with respect to functional outcomes (PCCC ≥ 2) and seizure coding (Fig. 4). The adjusted mean duration of mechanical ventilation was higher in the late DC (15 (95% CI: 13.0-17.0)) compared to 11.5 (95% CI: 9.4-13.5) in the early DC group (See eFigure 2). The sensitivity analysis showed higher odds for death aOR: 1.75 (95%CI: 0.96-3.22) in early (≤ 1 hour time to DC after admission) versus late DC (> 1 hour) groups.

**Fig. 4.**
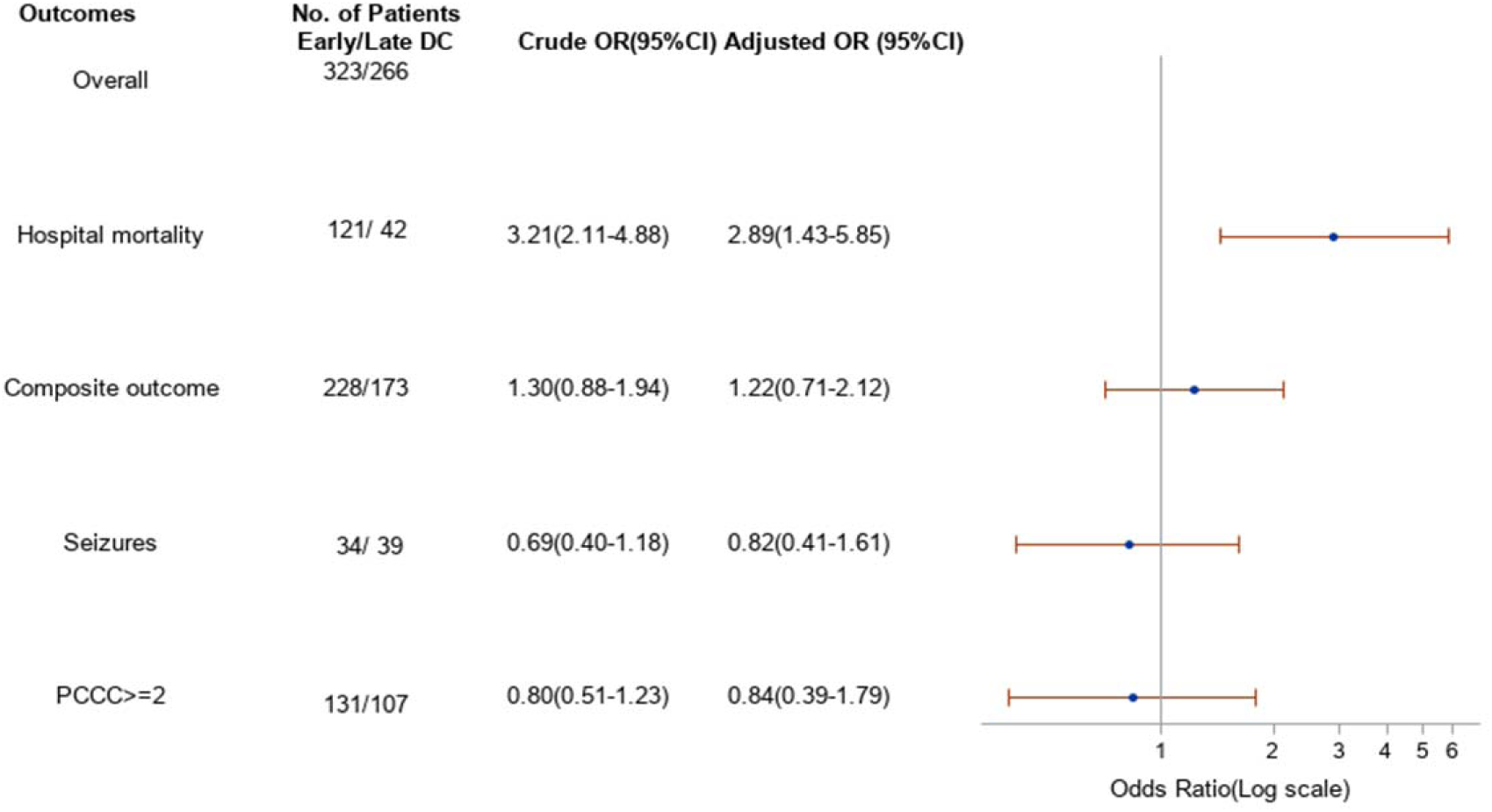
Odds ratio of adverse outcomes for early versus late decompressive craniectomy in children with severe traumatic brain injury in Germany 2016-2022. Adjustment variables: age, sex, single ICISS, coma, POFI, ICP monitoring and type of injury. Composite outcome = in-hospital death or PCCC ≥2

## Discussion

The indication for DC and its optimum timing to control ICP and prevent brain herniation in severe pediatric TBI remain undefined in medical practice guidelines, leaving the decision at the treating team’s discretion. This nationwide retrospective cohort study found that early DC was associated with higher in-hospital mortality, though survivors had fewer days on mechanical ventilation and shorter hospital stays compared to the late DC group. No clear difference in functional outcomes was observed between groups.

Previous RCTs, such as the DECRA and RESCUEicp trials, have evaluated outcomes after secondary DC in adult populations. In children, one RCT, several case series, and retrospective studies have evaluated the effect of DC on outcomes [3, 7, 8, 10, 20-24]. However, these studies often had limited sample sizes, or compared DC to medical management and had high risk of selection bias. A very limited number of pediatric TBI patients in retrospective studies has been compared regarding the timing effect of DC with varying definitions of early and late surgery time differences [7, 10, 25]. Our study, the first large-scale nationwide analysis on timing of DC in children and its association with outcomes, found that most DCs were performed on the first day following admission, with half occurring within two hours of hospital admission.

Early DC performed parallel to mass lesion evacuation, without prior ICP monitoring, was conducted within the first two hours after admission, and associated with more severe extracranial injuries in our study. In previous studies, primary DC as a prophylactic measure to prevent rapid ICP elevation post-mass lesion evacuation, generally involved patients with more severe TBI, including lower Glasgow Coma Scale scores, thicker hematomas, greater midline shifts, and more significant extracranial injuries [26-29]. In contrast, late DC was more frequently associated with prior ICP monitoring and the use of EVD indicating a more measured approach based on the progression of symptoms and ICP levels. Despite guidelines recommending ICP monitoring to manage ICP levels in sTBI, only half of the patients in our study received this continuous evaluation, which is particularly important in the late DC cases, where elevated ICP can prompt DC [30-32]. While there is no class I evidence regarding ICP monitoring after primary DC, several retrospective studies in adults concluded its usefulness in guiding therapy postoperatively, especially in the presence of brain swelling, and to reduce in-hospital mortality [33-35].

The variability in timing and population characteristics across studies evaluating the effect of DC timing in children or adults with sTBI makes comparison difficult. In our study, early DC was associated with higher lethality and shorter time to death, with a marked percentage of children undergoing hematoma evacuation during the same surgery. Conversely, the DECRA trial, which defined early DC as less than 38 hours and refractory to first-tier treatment, excluded patients with intracranial hemorrhage and reported equal mortality rates between early DC and MM [5]. In the RESCUEicp study, where DC was considered a last-tier therapy with a median time to randomization of 44.3 hours, demonstrated lower mortality rates. A recent retrospective study by Nagy et al. also found that the acute intervention group in children (defined as within 24 hours of admission) had higher mortality compared to the subacute group with no differences in functional outcomes (33.3% vs. 18.2%, respectively) [10]. However, multivariable analysis and thus adjustment for confounders were not performed due to the limited sample size. Two other retrospective studies have evaluated the association between timing and outcomes in adults (defining early surgery as 3- and 4-hours from injury to surgery respectively) and showed no differences in mortality compared to the late DC group [36, 37]. Another retrospective study that evaluated the association between early DC (time from injury to surgery less than 12 hours) and 14 days in-hospital mortality found that children with higher ISS had increased odds of death, however there were no comparison to a late DC group [22]. In our study, the increased lethality and rapid progression to death observed in the early DC group may reflect the severity of the initial injury rather than the impact of the intervention itself. However, survivors of early DC had shorter hospital stays and fewer days on mechanical ventilation compared to late DC, suggesting a more rapid and less complicated recovery.

The present study has several limitations, including lack of information on clinical reasoning for DC timing, physiological parameters from the shock room and ICU, and medical management before and after surgery. Additionally, time-related information such as duration from the accident to hospital admission are not available. There is potential for confounding by indication related to injury severity and the factors leading to hematoma evacuation with subsequent DC, as documentation on these factors may be lacking. However, we attempted to adjust for available confounders to the best of our ability, recognizing that often a subset of confounders can effectively mitigate much of the residual confounding. Future research should include prospective data collection and standardized criteria for surgical decision-making to optimize DC timing in pediatric sTBI. Despite these limitations, this study includes a considerable number of DC cases in children, providing a comprehensive description of DC timing and associated outcomes after sTBI in Germany.

Primary versus secondary DC are two different entities with their own clinical characteristics, timing, and indication. Early DC is often necessitated during the initial evacuation of intracranial mass lesions, but its association with higher lethality emphasizes the need for careful patient selection and individualized treatment strategies. A comprehensive risk assessment should guide clinical decision-making, weighing the potential long-term complications against this strategy. Potential complications, such as infections, cerebrospinal fluid leaks, and the need for re-surgery, must be discussed with parents to ensure informed decision-making regarding treatment strategy.

## Conclusion

This nationwide study of DC after severe pediatric TBI in Germany found that early DC, often necessary to evacuate a massive lesion, was associated with higher in-hospital mortality. Future research should focus on further developing timeline guidelines and incorporating standardized decision criteria to improve patient outcomes.

## Acknowledgment

None

## Research data statement

The data analyzed in this study is subject to the following licenses/restrictions: the original dataset can be accessed after inquiry to the Federal Bureau of Statistics of Germany. Requests to access these datasets should be directed to https://www.forschungsdatenzentrum.de/de.

## Supplementary

*eTable 1: Extracted ICD- and OPS-Codes and newly calculated variables.*

*eTable 2: Calculation of Pediatric complex chronic conditions classification.*

*eFigure 1: Directed acyclic graph to identify the minimally sufficient adjustment set for multivariable analyses.*

*eFigure 2: Adjusted least square means of mechanical ventilation in days(a), length of stay (b) and PCCC (c) by early versus late DC survivors’ cases. Adjusted model controlled for age, sex, single ICISS, coma, ICP monitoring, POFI and type of injury. Vertical bars denote 95% confidence intervals.*

