## Supplementary figures and images for "Timing of decompressive craniectomy and in-hospital mortality in children following severe traumatic brain injury using the German Hospital Database" (from another version of this preprint: version 1, 2024-11-21)

### eFig1

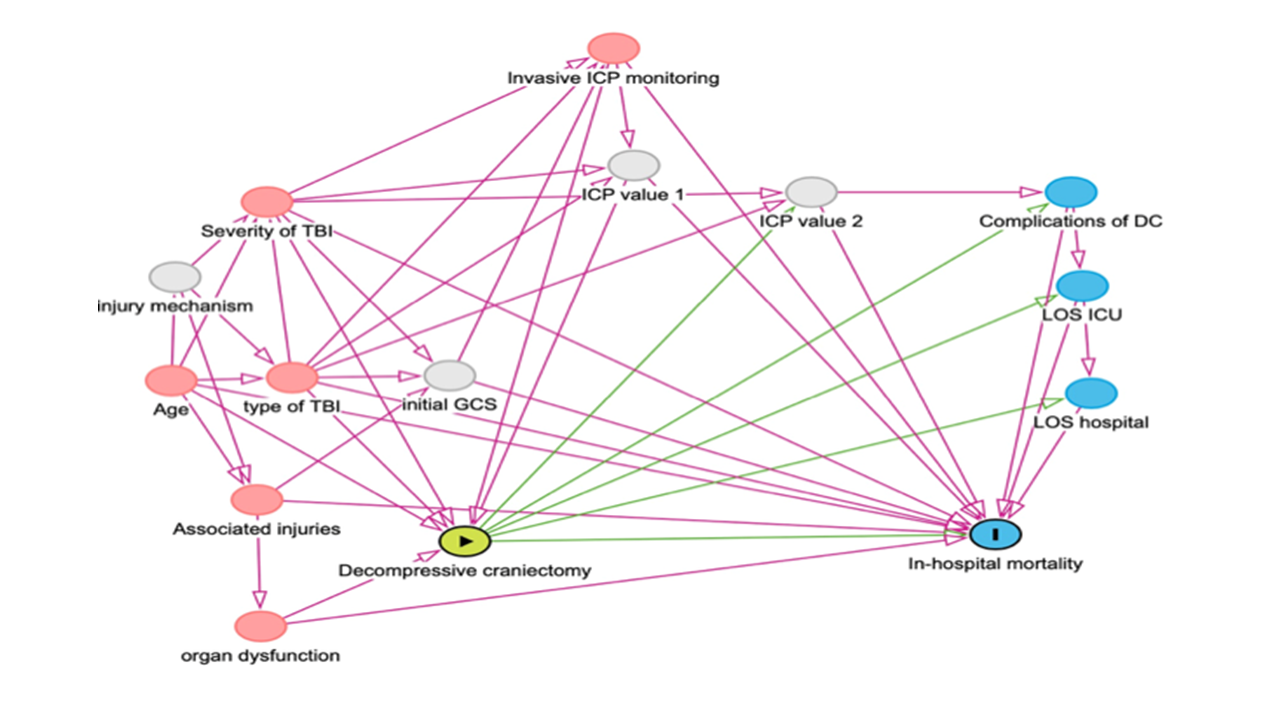

### eFig2

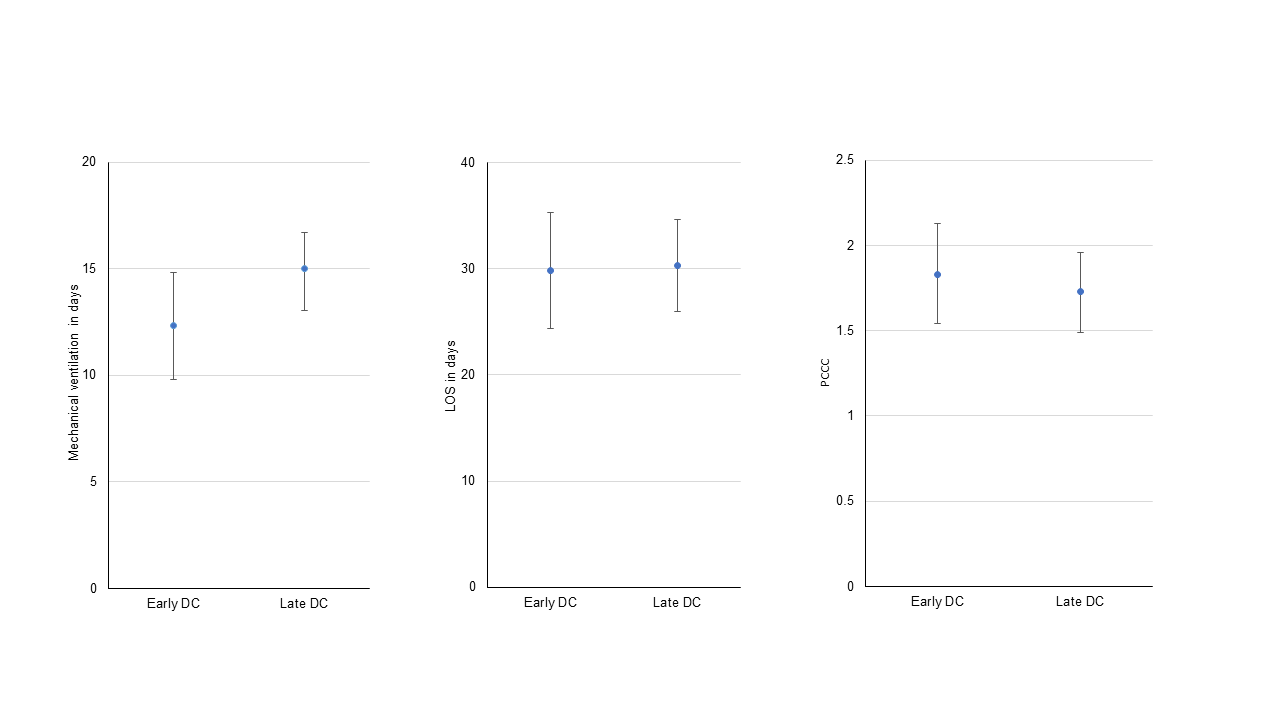
