## Supplementary material for "Timing of decompressive craniectomy and in-hospital mortality in children following severe traumatic brain injury using the German Hospital Database": eTable1 (from another version of this preprint: version 1, 2024-11-21)

| **Diagnoses** | **ICD-10-Code(s)** |
| --- | --- |
| Traumatic Brain injury | S06 (primary diagnosis) |
| Type of head injury |  |
| Brain oedema | S061 |
| Epidural Hemorrhage | S064 |
| Subdural Hemorrhage | S065 |
| Subarachnoid Hemorrhage | S066 |
| Complications |  |
| IHCA | I46 |
| OHCA | U6913 |
| Seizures | G40 |
| Epileptic state | G41 |
| Coma | R402,S0672,S0673 |
| DIC | D651 |
| Organ failure |  |
| Cardiovascular (NOF_kard) | R57, P29 |
| Pulmonary (NOF_pulm) | P28, P22.0, P25, P27, |
| Coagulation (NOF_coag) | P60, P61.0 |
| Renal (NOF_ren) | N17 |
| Hepatic (NOF_hep) | K72, R740 |
| Neurologic (NOF_CNS) | R40 |
| Neonatal organ failure score | Sum of binary scores NOF_kard, NOF_pulm, NOF_ren, NOF_coag, NOF_GI, NOF_hep, NOF_CNS, Dialysis, ECMO [continuous] |

**Supplementary table 1:** Extracted ICD- and OPS-Codes and newly calculated variables.

| **Procedures** | **OPS-Codes** |
| --- | --- |
| Decompressive craniectomy | 50120,50101,50104 |
| Evacuation of hematoma | 50131,50134,50122 |
| ICP monitoring | 50291 |
| EVD | 50220 |
| ECMO | 8852 |
| **Other variables extracted from the basic data set** | |
| Sex | Included in the DRG data set |
| Length of stay (days) | Included in the DRG data set |
| Ventilation (hours) | Included in the DRG data set |
| Death | Reason for discharge = 7 |
| Early transfer | Reason for discharge = 6 or 8 and duration of hospital stay < 24 hours |
