## Supplementary material for "Timing of decompressive craniectomy and in-hospital mortality in children following severe traumatic brain injury using the German Hospital Database": eTable1 (from another version of this preprint: version 1, 2024-11-21)

**Supplementary table 2:** Calculation of Pediatric complex chronic conditions classification

| PCCC subscore | **ICD-10-Code(s)** |
| --- | --- |
| Neurological (CCC_neuro) | Q00, Q01, Q02, Q03, Q04, Q05, Q06, Q07, G901, F71, F72, F73, E75, F842, G11, G12, G25.3, G31.1, G31.8, G31.9, G32.8, G91.1, G93.8, G93.9, G94, G95.18, G95.88, G90.9, Q85.1, G80, G40, G37.1, G37.2, G37.8, G81, G82, G83.5, G83.9, G93.1, G93.5, R40, I63, G71, G72, G10, G20, G21.1, G21.8, G23.0, G23.1, G23.2, G24.0, G24.8, G25.3, G25.4, G25.5, G25.8, G25.9, T85.0, T85.1, T85.7, Z98.2, Z99 |
| Cardiological (CCC_cardio) | Q20, Q21.2, Q21.3, Q21.4, Q21.8, Q21.9, Q22, Q23, Q24, Q25.1, Q25.2, Q25.3, Q25.5, Q25.6, Q25.7, Q25.8, Q25.9, Q26, Q28.2, Q28.3, Q28.9, I34.0, I34.8, I36.0, I36.8, I37.0, I37.8, I42, I43, I51.5, I44, I45, I47, I48, I49, R00.1, I27, I50.9, I51.7, I51.8, I63.1, I63.2, Z95.1, T82.5, T82.1, T82.0, T82.2, T82.6, T82.7, Z95.0, Z95.2, Z95.3, Z95.8, Z95.9, Z45.01, Z45.02, Z45.09, T86.2, Z94.1 |
| Respiratory (CCC_resp) | Q30, Q31, Q32, Q33, Q34, P28.0, G47.32, I27.8, I43, J84.1, J96, Z90.2, E84, J95.0, J95.8, Z43.0, Z93.0, Z99.0, Z99.1, T86.81, Z94.2 |
| Renal (CCC_renal) | Q60, Q61, Q62, Q63, Q64, N18, Z90.5, Z90.6, G83.4, N31.2, N31.9, T85.71, Z93.5, Z93.6, Z91.1, Z99.2, Z43.5, Z43.6, Z46.6, T86.1, Z94.0 |
| Gastroenterological (CCC_gastro) | Q39.0, Q39.1, Q39.2, Q39.3, Q39.4, Q41, Q42, Q43, Q45, K73, K74, K75, K76.0, K76.1, K76.2, K76.3, K76.5, K76.8, K50, K51, I82.0, K55.1, K56.2, K59.3, Z98.0, Z90.3, Z90.4, Z93.1, Z93.2, Z93.3, Z93.4, Z43.1, Z43.2, Z43.3, Z43.4, T86.4, T86.8, Z94.4, Z94.88 |
| Hematological/immunological (CCC_hema_immu) | D55, D56, D57, D58, D60, D61, D71, D8, D72.0, M30.3, M35.9, D66, D68.2, D69.4, D69.3, D70.0, D70.5, D76.1, D76.2, D76.3, D86.9, B20, B21, B22, B23, B24, M30.0, M31.0, M31.3, M31.4, M31.6, M32.1, M33.9, M34.0, M43.1, M34.9 |
| Metabolic (CCC_metab) | E70.0, E70.2, E70.3, E70.8, E71, E72.0, E72.1, E72.3, E72.4, E72.8, E72.9, E74, E75, E77.0, E77.1, E78, E88.8, E76.0, E76.1, E76.2, E76.3, E85, E78.6, E79.1, E79.8, E80.4, E80.5, E80.6, E80.7, E93.0, E83.1, E83.3, E83.4, D84.1, E88, H49.8, E00.9, E23.0, E23.2, E23.3, E23.7, E24.0, E24.2, E24.3, E24.8, E24.9, E26.8, E25.0, E25.8, E25.9, Z46.8, Z96.4 |
| Genetic malformations (CCC_genet_malform) | Q90.9, Q91.3, Q91.4, Q91.7, Q92.8, Q93, Q95.0, Q96.9, Q97, Q98, Q99.8, Q99.9, E34.3, M41.0, M41.2, M41.39, M41.8, M41.9, M43.3, M96.5, Q72.2, Q75.0, Q75.2, Q75.9, Q76.0, Q76.1, Q76.2, Q76.4, Q76.5, Q76.6, Q77, Q78.0, Q87.1, Q78.2, Q78.3, Q78.4, Q78.8, Q78.9, K44.9, Q79.0, Q79.1, Q79.2, Q79.3, Q79.4, Q79.5, Q79.9, Q81, Q87.1, Q87.2, Q87.3, Q87.4, Q87.8, Q89.7, Q89.9, Q99.2 |
| Malignancy (CCC_malign) | C, D0, E34.0, D37, D38, D39, D4, Q85.0, T86.0, Z94.80, Z84.81 |
| Neonatal (CCC_neo) | P05, P07.0, P07.2, P10.0, P10.1, P10.4, P52.4, P52.8, P11.5, P21.0, P21.9, P96, P25, P27.0, P27.1, P27.8, P91.6, P35.0, P35.1, P35.2, P56.0, P57.0, P57.8, P61.3, P61.4, P77, P83.2, P91.2 |
| Dependency from technical devices (CCC_tech_dep) | T84.01, T84.02, T84.03, T84.04, T84.05, T84.06, T84.08, T84.11, T84.12, T84.18, T84.4, T84.5, T84.6, T84.7, T86.9, T86.0, T86.8, T86.5, T87.0, T87.1, Z99.8, Z93.1, Z93.2, Z93.3, Z93.4, Z43.1, Z43.2, Z43.3, Z43.4, T85.71, Z93.5, Z93.6, Z91.1, Z99.2, Z43.5, Z43.6, Z46.6, J95.0, J95.8, Z43.0, Z93.0, Z99.0, Z99.1, T82.5, T82.1, T82.0, T82.2, T82.6, T82.7, Z95.0, Z95.2, Z95.3, Z95.8, Z95.9, Z45.01, Z45.02, Z45.09, T85.0, T85.1, T85.7, Z98.2, Z99 |
| Transplantation recipient* (CCC_transplant) | Z94, T86 |
| PCCC score | Sum of binary sores CCC_neuro, CCC_cardio, CCC_resp, CCC_renal, CCC_gastro, CCC_hema_immu, CCC_metab, CCC_genet_malform, CCC_malign, CCC_neo, CCC_tech_dep, CCC_transplant |

* solid organ or bone marrow transplant
